# Disentangling the causal effects of education and participation bias on Alzheimer’s disease using Mendelian Randomization

**DOI:** 10.1101/2024.07.09.24310096

**Authors:** Aadrita Chatterjee, Clémence Cavaillès, Neil M Davies, Kristine Yaffe, Shea J Andrews

**Affiliations:** Department of Psychiatry and Behavioral Sciences, University of California, San Francisco, CA, USA; San Francisco Veterans Affairs Health Care System, San Francisco, CA, USA; Division of Psychiatry, University College London, Maple House, 149 Tottenham Court Rd, London W1T 7NF; Department of Statistical Science, University College London, London WC1E 6BT, UK; K.G. Jebsen Center for Genetic Epidemiology, Department of Public Health and Nursing, Norwegian University of Science and Technology, Norway

**Keywords:** education, participation bias, Alzheimer’s disease, Mendelian Randomization, causal inference

## Abstract

**Introduction:** People with university degrees have a lower incidence of Alzheimer’s Disease (AD). However, the relationship between education and AD could be due to selection, collider, and ascertainment biases, such as lower familiarity with cognitive testing or the fact that those with degrees are more likely to participate in research. Here, we use two-sample Mendelian randomization (MR) to investigate the causal relationships between education, participation, and AD.

**Method:** We used genome-wide association study (GWAS) summary statistics for educational attainment, three different measures of participation, AD (clinically diagnosed AD), and AD/ADRD (clinical diagnosis and family history of AD and related dementias). Independent genome-wide significant single nucleotide polymorphisms (SNPs) were extracted from the exposure summary statistics and harmonized with the outcome SNPs. Fixed-effects inverse variance weighted meta-analysis was the primary MR method; Cochran’s Q statistic and MR Egger intercept were used to test for heterogeneity and pleiotropy, and Radial-MR was used to identify outliers. Sensitivity analyses included MR Egger, Weighted Median, and Weighted mode. Bidirectional analyses were used to test if AD pathology affects participation and multivariable MR (MVMR) assessed independent exposure-outcome effects.

**Results:** Educational attainment reduced the risk of AD (OR_IVW_ 95% CI= 0.70 [0.63, 0.79], p = 8e-10), and the results were robust based on sensitivity analyses. However, education increased the risk of AD/ADRD, though the results were not robust to sensitivity analyses (OR_IVW_ 95% CI= 1.09 [1.02, 1.15], p = 0.006). Participation in MHQ reduced the odds of AD (OR_IVW_ 95% CI= 0.325 [0.128, 0.326], p = 0.01). When adjusting for participation in MVMR, education remained associated with a reduced risk of AD (OR_IVW_ 95% CI= 0.76 [0.62, 0.92], p = 0.006).

**Conclusion:** Univariate MR analyses indicated that education and participation reduced the risk of AD. However, MR also suggested that education increased the risk of AD/ADRD, highlighting the inconsistencies between clinical and proxy diagnoses of AD, as proxy-AD may be affected by selection, collider, or ascertainment bias. MVMR indicated that participation is unlikely to explain the effect of education on AD identified in MR, and the protective effect of educational attainment may be due to other biological mechanisms, such as cognitive reserve.

## Introduction

Epidemiological studies have found that people with higher educational attainment have a lower incidence of Alzheimer’s Disease (AD). These associations may be due to cognitive reserve, whereby education reduces the impact of neuropathologic lesions in the brain (1,2). Increased educational opportunities and improved cardiovascular health have also been linked to the observed secular decline in the incidence of dementia (3,4). However, the observed protective effect of education on AD may be influenced by participation bias, as individuals with more education are more likely to participate in research studies (5). In a study examining participation in clinical research and education, individuals with higher levels of education, such as college students, exhibited a greater likelihood of participation in clinical trials (6–8). In comparing the National Alzheimer’s Coordinating Center sample (NACC) Alzheimer’s Disease Research Centers to the nationally representative Health and Retirement Study (HRS), NACC participants had higher levels of education compared to HRS participants, further supporting a positive relationship with education and participation in clinical studies (9).

Selection bias, collider bias, and participation bias can all influence the observed relationship between education and AD. Selection bias occurs when the participants in a study are not representative of the general population (10). In collider bias, researchers select their data based on a variable that is influenced by both the exposure and outcome they are studying. Participation bias represents a form of collider and selection bias because any observed association between education and AD may reflect the combined influence of education and participation bias and could lead to distorted causal interpretations (Supplementary Figure 1) (10). For example, selective participation could create a spurious association between higher education and lower prevalence of AD within the study sample. This bias occurs not because education directly influences AD risk, but because of the non-random selection process based on both education level and AD status.

The observed effects of education in Mendelian randomization studies could be due to either the causal effects of education or participation bias. This is important because if the effects are due to collider bias, then educational attainment is unlikely to be an effective modifiable risk factor for Alzheimer’s disease, and recent secular changes in educational attainment are unlikely to affect rates of disease in the population.

Mendelian Randomization uses instrumental variables —genetic variants associated with the exposure of interest— to investigate the causal effect of an exposure on an outcome (11). Due to the random inheritance of alleles at conception and meiosis, conditional on parental genotype, genetic variants associated with exposure cannot be confounded (12). Furthermore, reverse causation is impossible, as the environment cannot affect germline genetic variation post-conception. Multivariable MR (MVMR) extends this approach by using genetic variants associated with multiple exposures as instruments to estimate the “direct” and indirect effects of each exposure on the outcome (12). MVMR analyses have shown a bidirectional effect between education and cognitive ability; when examining the total effects of education on AD, it was found that cognitive abilities mediates the impact of education on AD (13). Additionally, cortical surface area, volume, and intrinsic curvature were found to be associated with educational attainment (14).

To date, the effect of participation on the protective effect of educational attainment on AD needs to be further investigated. The effect of education and AD can influence participation (and subsequentially selection) therefore MR can be employed to overcome certain biases as the impacts of selection bias are likely to be less than other biases such as pleiotropy (15). This study used genetic correlations and bidirectional and multivariable two-sample Mendelian randomization (MR) to investigate the causal relationships between education, participation, and AD and determine if education’s effect on AD is due to participation bias.

## Methods

### Data Sources

Genome-wide association study (GWAS) summary statistics were obtained for each exposure and outcome dataset (Table 1 and Supplementary Table S1). The GWAS for education measured educational attainment as the number of years of schooling (n= 3,037,499, n_loci_ = 3,952) (16). For participation, we used one primary measure of participation involving participation in an optional mental health questionnaire (MHQ) of the UK Biobank (n= 451,036, n_loci_ = 32) as it was the most statistically powered (17). Two other GWASs’ of participation were used to validate our results across different measures and included a weighted GWAS based on a probability model that has individual participation probabilities as the outcome (n_effective_ = 102,215, n_loci_ = 28) (18). This model adjusted for nonresponse by giving greater weight to overrepresented and underrepresented individuals, thus creating a more representative pseudo-population that mimics the Health Survey England, which was used as the reference sample. The next measure of participation used estimated factor scores for the general “I don’t know” behavior across UK Biobank survey questions (n= 360,628, n_loci_ = 35) (19). Two GWASs’ were used for AD: the first involved clinically diagnosed AD cases (n= 94,437, n_loci_ = 25 LOAD risk loci), which we refer to as “AD” (20). The second leveraged clinical case-control series, in addition to self-reported family history of dementia, to conduct a GWAS-by-proxy (GWAX) (n = 788,989, n_loci_ = 72) of AD and related dementias which we refer to as “AD/ADRD” (21). All cohorts included age and sex as covariates, and all individuals were of European ancestry. All GWAS’s were standardized using ‖MungeSumstats’ version 1.10.1 (22).

**Table 1.** Summary of GWASs used in study.

| **Study** | **Trait** | **Cohort/**<br>**consortium** | **N** | **Number**<br>**of loci** | **Ancestry** |
| --- | --- | --- | --- | --- | --- |
| Kunkle et al,<br>2019 | Late-onset<br>Alzheimer's<br>disease | IGAP | 94,437 | 25 | European |
| Bellenguez et<br>al, 2022 | Proxy AD | EADB<br>consortium and<br>the UK<br>Biobank | 487,511 | 72 | European |
| Okbay et al,<br>2022 | Education | UK Biobank | 3,037,499 | 3,952 | European |
| Tyrrell et al,<br>2021 | Participation in<br>MHQ | UK Biobank | 294,787 | 32 | European |
| Schoeler et al,<br>2021 | Weighted<br>participation | UK Biobank,<br>HSE, UK<br>Census | 94,643 –<br>102,215 | 28 | European |
| Mignogna et<br>al, 2023 | Nonresponse<br>participation | UK Biobank,<br>Add Health | 360,628 | 35 | European |

IGAP, International Genomics of Alzheimer's Project; HSE, Health Survey England; Add Health, National Longitudinal Study of Adolescent to Adult Health

### Genetic correlations

We estimated genetic correlations between each trait using Linkage Disequilibrium Score Regression (LDSC) implemented by GenomicSEM v0.0.5 (23). Genetic correlations quantify the degree of shared genetic influence between two traits, representing the proportion of variance in these traits that can be attributed to common genetic influences (24). LDSC quantifies heritability by assessing the correlation between genetic variants across the genome and the trait of interest, using a European reference panel from 1000 Genomes to estimate linkage disequilibrium (LD) (24).

### Mendelian Randomization

#### Selection of genetic instruments and data harmonization

We first performed clumping (r^2^ = 0.001, 10Mb clumping window, EUR reference) to identify and retain independent genome-wide significant (p < 5e-8) SNPs using the OpenGWAS API.

For instrumental variables missing from the outcome GWAS dataset, LD proxies were identified using LDlinkR version 5 (reference = EUR; r^2^ > 0.8) (25). The exposure and outcome datasets were harmonized to ensure that their SNP effects corresponded to the same effect allele with palindromic variants inferred using their allele frequencies (26,27). The *APOE* region (19:44912079-19:45912079, build 37) was removed due to its known pleiotropic effects (28).

#### Statistical analysis

MR is a statistical technique that leverages genetic variants as instrumental variables to estimate causal relationships between an exposure and an outcome. MR relies on the principle that genetic variants are randomly assigned at conception and thus free from confounding, making them ideal instruments for causal inference. MR holds three key assumptions: the genetic variants associate with the exposure; there are no controlled confounders of the genetic variant-outcome association; all the effects of the genetic variant on the outcome are mediated via the exposure of interest (12).

We performed univariate MR to estimate the causal effect of education on each participation measure and AD. We also estimated the effect of genetic liability to AD on education and each participation measure. Fixed effects Inverse-variance weighted (IVW) was the primary analysis method as it is the most precise. Fixed effects IVW weighs variant-exposure and variant-outcome associations by the inverse of their variances to provide a single causal estimate, assuming that all instruments are valid and there is no horizontal pleiotropy (29).

#### Diagnostics

To test the validity of the instrumental variable assumptions, we used F-statistics to assess the strength of the genetic instruments, Cochran’s Q test for heterogeneity, and the MR Egger regression intercept for horizontal pleiotropy (12). Higher F-statistics (F statistic >10) indicate stronger instruments and are less likely to lead to weak instrument bias (12). Radial MR version 1.0 was used to detect outliers that were omitted from the MR analysis (30).

#### Sensitivity Analyses

For each MR analysis, sensitivity analyses were performed to test the robustness of the causal association between the exposure and outcome in either heterogeneity or horizontal pleiotropy. These include MR Egger, Weighted Median (WME), and Weighted Mode Based Estimator (WMBE), with each method having different assumptions (31–33). The assumption of no horizontal pleiotropy is relaxed in MR Egger; WME combines multiple genetic instruments to estimate causal effects and provides valid results even when at least 50% of the instruments are valid; WMBE is unbiased when the modal estimate across the SNPs is from a valid (i.e. non-pleiotropic) SNP (12). We interpreted our results as robust evidence of a causal effect when the IVW analysis was significant (p<0.05) after outlier removal and where there was no evidence of heterogeneity (p>0.05) or pleiotropy (p>0.05). In the presence of heterogeneity or pleiotropy, robust causal associations were those where at least one of the sensitivity analyses was also significant (p<0.05) and had the same direction of effect.

#### Multivariable Mendelian Randomization

Multivariable Mendelian randomization is an extension of univariate MR that includes instrumental variables associated with multiple exposures to be included in the analysis. Therefore, multivariable MR allows us to evaluate the direct causal effect of an exposure on an outcome, while univariate MR only estimates the total causal effect (12). The two exposure/outcome datasets were harmonized using TwoSampleMR version 0.5.11. We performed multivariable MR and evaluated the results using diagnostics and sensitivity analyses to determine if the effect of education on AD was being mediated by participation bias using the MVMR package version 0.3 and MendelianRandomization package version 0.9.0 (34)(35). A robust causal effect in the MVMR analysis is similarly defined as the univariate analysis.

All statistical analyses were carried out using R version 4.3.0. The code used to conduct the analyses is available at: https://github.com/AndrewsLabUCSF/Aadrita-AD-participation-education.

## Results

### Genetic correlations

We found medium-strong positive genetic correlations between the three participation measures (Figure 1). Education exhibited the strongest positive correlation with weighted participation, followed by participation and nonresponse participation. Education was also negatively correlated with AD. However, it was not correlated with AD/ADRD. In contrast, participation and both AD and AD/ADRD were weakly negatively correlated (Figure 1).

**Fig 1:**
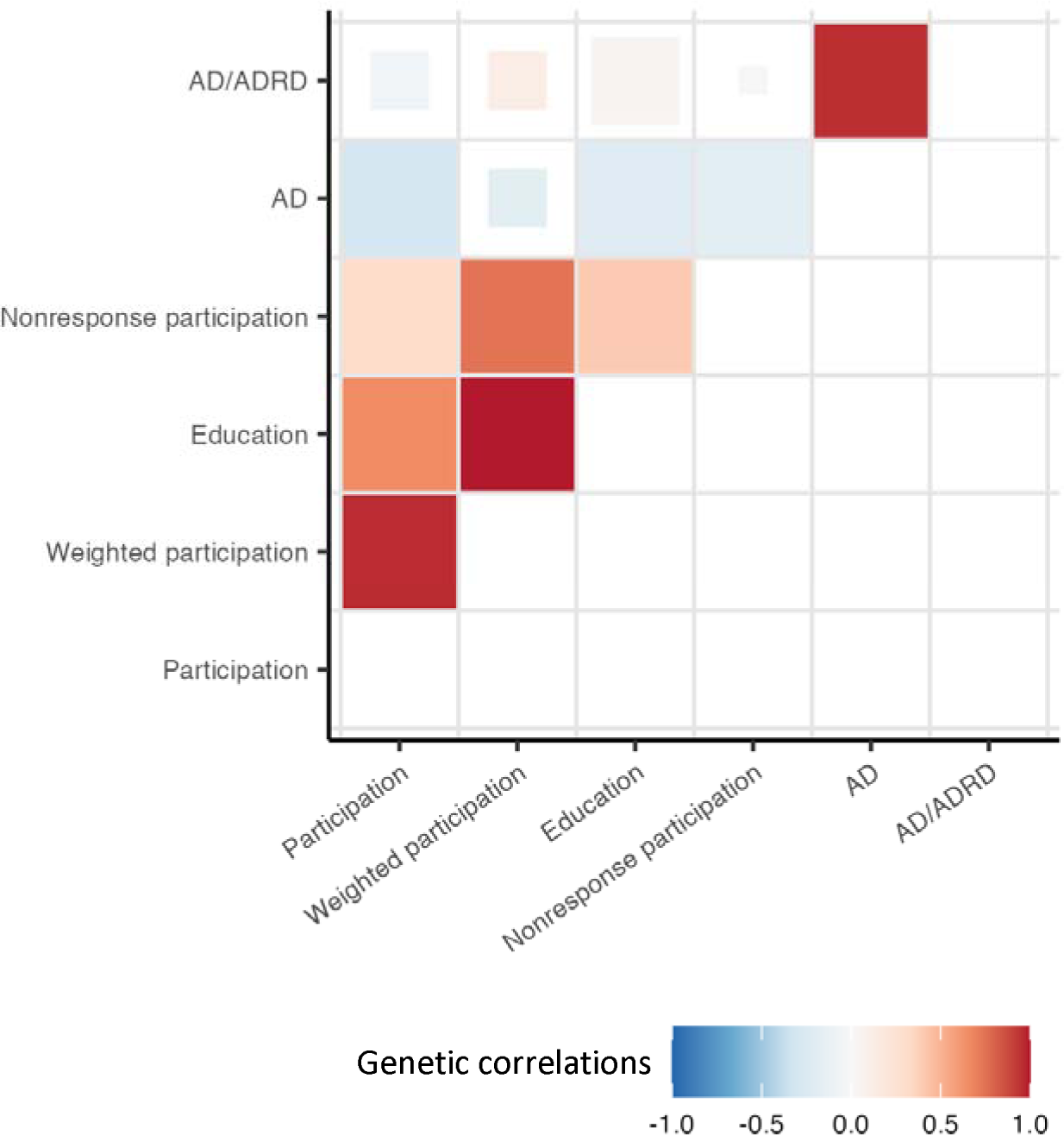
Genetic correlations of education, participation, weighted participation, nonresponse participation and AD and AD/ADRD. Genetic correlations between three different participation measures, education, AD and AD/ADRD. Blue indicates a negative correlation while red indicates a positive correlation. P values are represented by the size of the colored squares where larger squares represent a smaller P value. Genetic correlations were calculated using GWAS summary statistics from previous studies and LD score regression. Nonresponse participation is reverse coded.

### Higher Educational attainment is associated with a reduced risk of AD, but an increased risk of AD/ADRD

The Mendelian randomization estimates suggested that an additional year of education reduced the odds of AD across all sensitivity analyses (OR_IVW_ 95% CI= 0.7 [0.63, 0.79], p = 8e-10; Table 2, Figure 2 and Supplementary Fig 2A) but our results were not robust to heterogeneity and pleiotropy. In contrast, the Mendelian randomization estimates suggested that an additional year of education increased the odds of AD/ADRD (OR_IVW_ 95% CI= 1.09 [1.02, 1.15], p = 0.006; Table 3, Figure 3 and Supplementary Figure 3A). However, there was evidence of heterogeneity, and the sensitivity analyses were consistent with the null.

**Fig 2:**
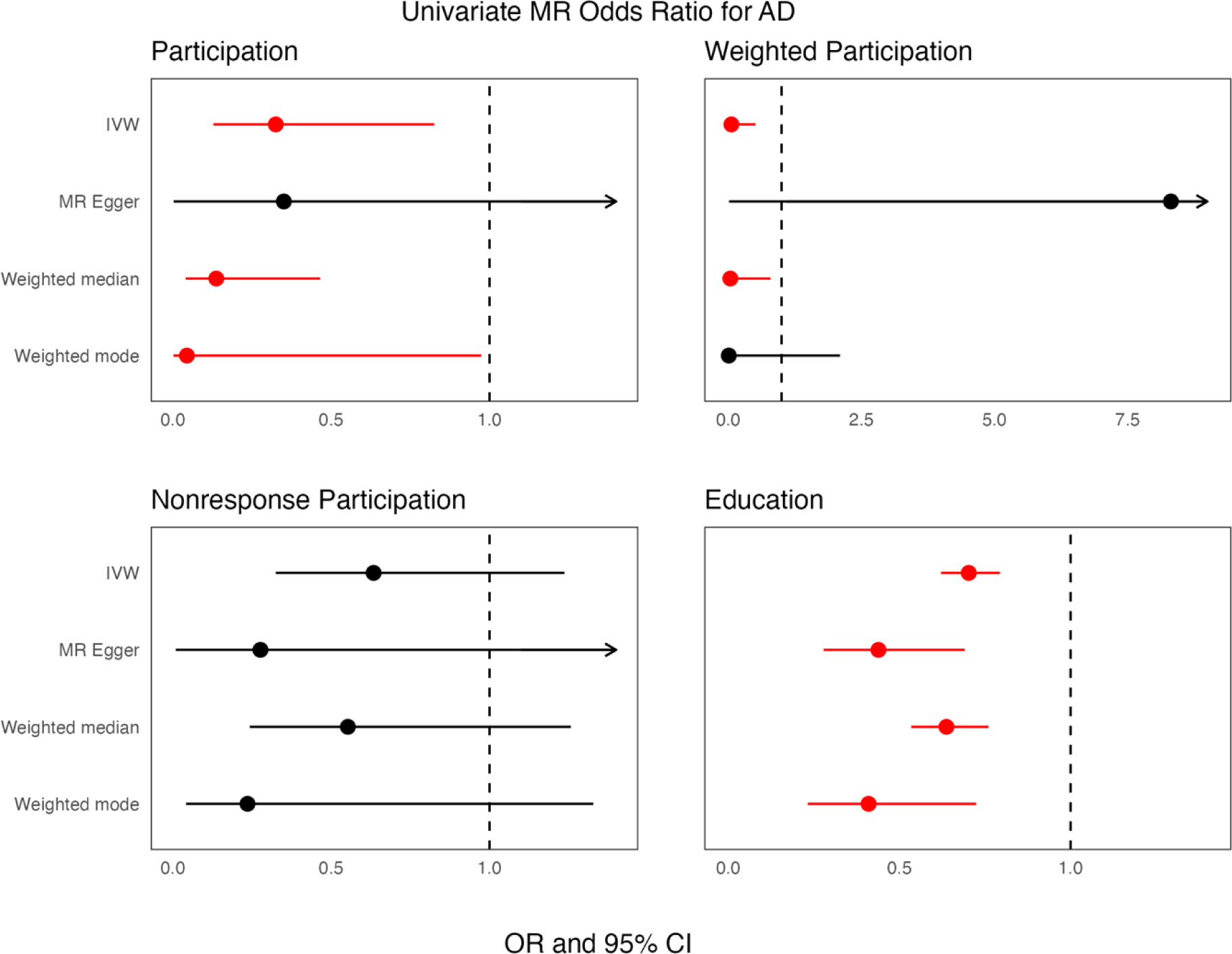
Univariate odds ratio of the association between participation measures, education and AD. Forest plots showing the univariate MR odds ratio for the four different traits (participation, weighted participation, nonresponse participation and education) and AD. Red represents significant results and arrows represent CI intervals that continue past the x axis.

**Fig 3:**
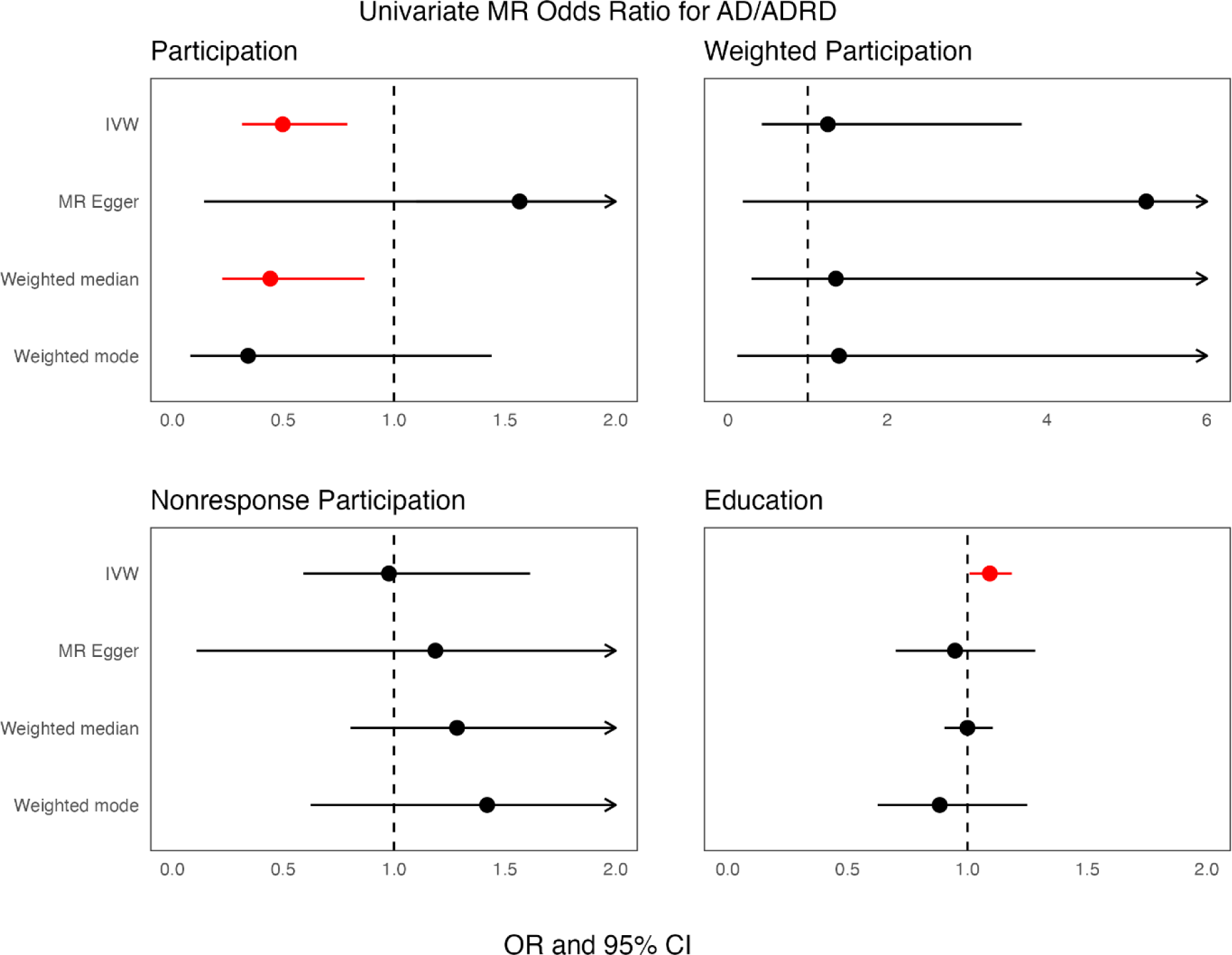
Univariate odds ratio of the association between participation measures, education and AD/ADRD. Forest plots showing the univariate MR odds ratio for the four different traits (participation, weighted participation, nonresponse participation and education) and AD/ADRD. Red represents significant results and arrows represent CI intervals that continue past the x axis.

**Table 2.** Summary of univariate MR results with AD.

|  |  | SN<br>P | F<br>stat<br>s | IVW |  | MR-Egger |  | WME |  | WMBE |  | Cochran's<br>Q test | MR-Egger<br>intercep<br>t |
| --- | --- | --- | --- | --- | --- | --- | --- | --- | --- | --- | --- | --- | --- |
| Exposure | Outcome |  |  | Beta<br>(se) | P<br>value | Beta<br>(se) | P value | Beta (se) | P value | Beta<br>(se) | P value | Q p value | P value |
| Education | AD | 464 | 55.<br>1 | -0.34<br>(0.06) | 8e-10 | -0.89<br>(0.23) | 1.2e-04 | -0.45<br>(0.08) | 9.6e-08 | -0.88<br>(0.31) | 0.005 | 2e-04 | 0.01 |
| Participation | AD | 36 | 34.<br>9 | -1.12<br>(0.47) | 0.01 | -1.04<br>(2.47) | 0.67 | -1.98<br>(0.62) | 0.001 | -3.11<br>(1.57) | 0.05 | 0.10 | 0.97 |
| Weighted<br>participation | AD | 18 | 31.<br>3 | -2.83<br>(1.1) | 0.01 | 2.11<br>(3.22) | 0.52 | -3.24<br>(1.5) | 0.03 | -4.64<br>(2.53) | 0.08 | 0.94 | 0.12 |
| Nonresponse<br>participation | AD | 29 | 39.<br>9 | -0.45<br>(0.34) | 0.18 | -1.28<br>(1.7) | 0.46 | -0.59<br>(0.4) | 0.1 | -1.44<br>(0.85) | 0.1 | 0.033 | 0.02 |
| **Reverse**<br>**causality** |  |  |  |  |  |  |  |  |  |  |  |  |  |
| AD | Education | 27 | 105<br>.2 | 0.004<br>(0.003) | 0.22 | 0.007<br>(0.005) | 0.22 | 0.01<br>(0.003) | 0.002 | 0.009<br>(0.002) | 0.003 | 1.71e-06 | 0.57 |
| AD | Participation | 28 | 107<br>.3 | -0.005<br>(0.001) | 4.7e-05 | -0.005<br>(0.002) | 0.014 | -0.002<br>(0.002) | 0.19 | -0.002<br>(0.002) | 0.32 | 0.056 | 0.71 |
| AD | Weighted<br>participation | 25 | 117<br>.19 | 7.5e-05<br>(0.001) | 0.95 | -3.9e-04<br>(0.001) | 0.84 | -6.14<br>(0.001) | 0.97 | 1.83<br>(0.001) | 0.99 | 0.28 | 0.73 |
| AD | Nonrespons<br>e<br>participation | 24 | 117<br>.19 | -0.004<br>(0.002) | 0.059 | -0.003<br>(0.003) | 0.27 | -0.005<br>(0.003) | 0.11 | -0.003<br>(0.003) | 0.26 | 0.1 | 0.98 |

**Table 3.**
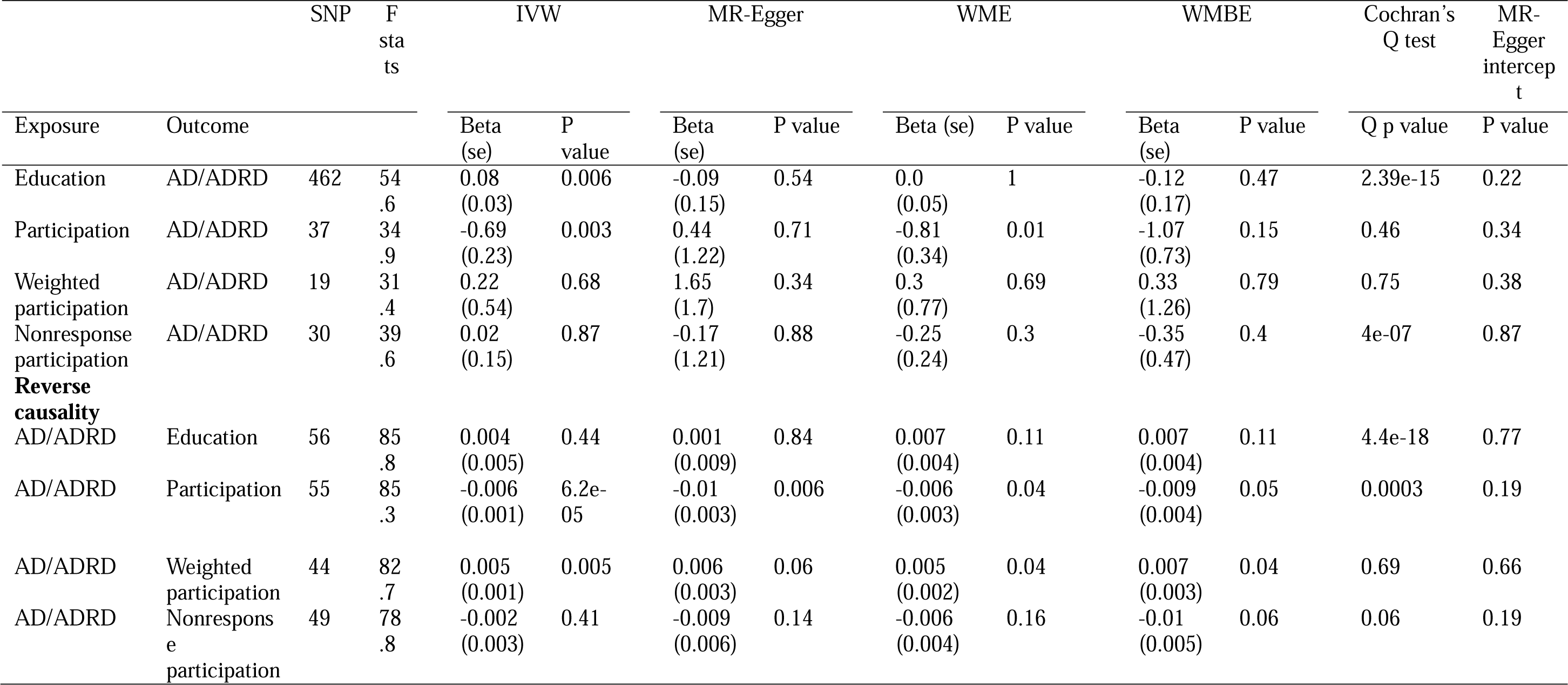
Summary of univariate MR results with AD/ADRD.

### Participation is associated with a reduced risk of AD

Participation in MHQ reduced the odds of AD (OR_IVW_ 95% CI= 0.325 [0.128, 0.326], p = 0.01; Table 2, Figure 2 and Supplementary Figure 2B) and was supported by similar results in the weighted participation measure (OR_IVW_ 95% CI= 0.0586 [0.0067, 0.510], p = 0.01; Table 2, Figure 2 and Supplementary Figure 2C). MHQ decreased the odds of AD/ADRD (OR_IVW_ 95% CI= 0.49 [0.31, 0.79], p = 0.003; Table 3, Figure 3 and Supplementary Figure 3B).

### Bidirectional analysis showed an effect of AD on participation but no effect of AD on education

Genetic liability for AD reduced the odds of participation in the MHQ (OR_IVW_ 95% CI= 0.994 [0.98, 0.99], p = 4.7e-05), and our results were robust to heterogeneity and pleiotropy (Table 2 and Supplementary Figure 4B). Similarly, genetic liability to AD/ADRD reduced the odds of participation (OR_IVW_ 95% CI= 0.993 [0.99, 0.996], p = 6.24e-05; Table 3 and Supplementary Figure 5B) and the results were robust. On the other hand, genetic liability to AD/ADRD increased the odds of weighted participation (OR_IVW_ 95% CI= 1.005 [1.001, 1.008], p = 0.005; Table 3 and Supplementary Figure 5C). There was little evidence that genetic liability for AD or AD/ADRD affected educational attainment.

### The protective effect of education on AD is not mediated by participation

When adjusting for participation in the MHQ in MVMR analysis, an additional year of education continued to reduce the odds of AD (OR_IVW_ 95% CI= 0.76 [0.62, 0.92], p = 0.006; Figure 4 and Supplementary Table S2). Instrument strength was low for education and participation (conditional F statistic for education= 3.41 and participation= 2.41; Figure 4 and Supplementary Table S2), suggestive of weak instruments. When adjusting for education in the MVMR, IVW estimates were non-significant for participation (OR_IVW_= 0.58, 95% CI= [0.22, 1.51], p = 0.27; Figure 4 and Supplementary Table S2) and this was supported by the weighted participation measure (OR_IVW_= 1.63, 95% CI= [0.39, 6.69], p = 4.9e-01; Figure 5 and Supplementary Table S2). Instrument strength was low for education and weighted participation (F statistic for education= 10.86 and weighted participation= 1.42), which is suggestive of weak instruments.

**Fig 4:**
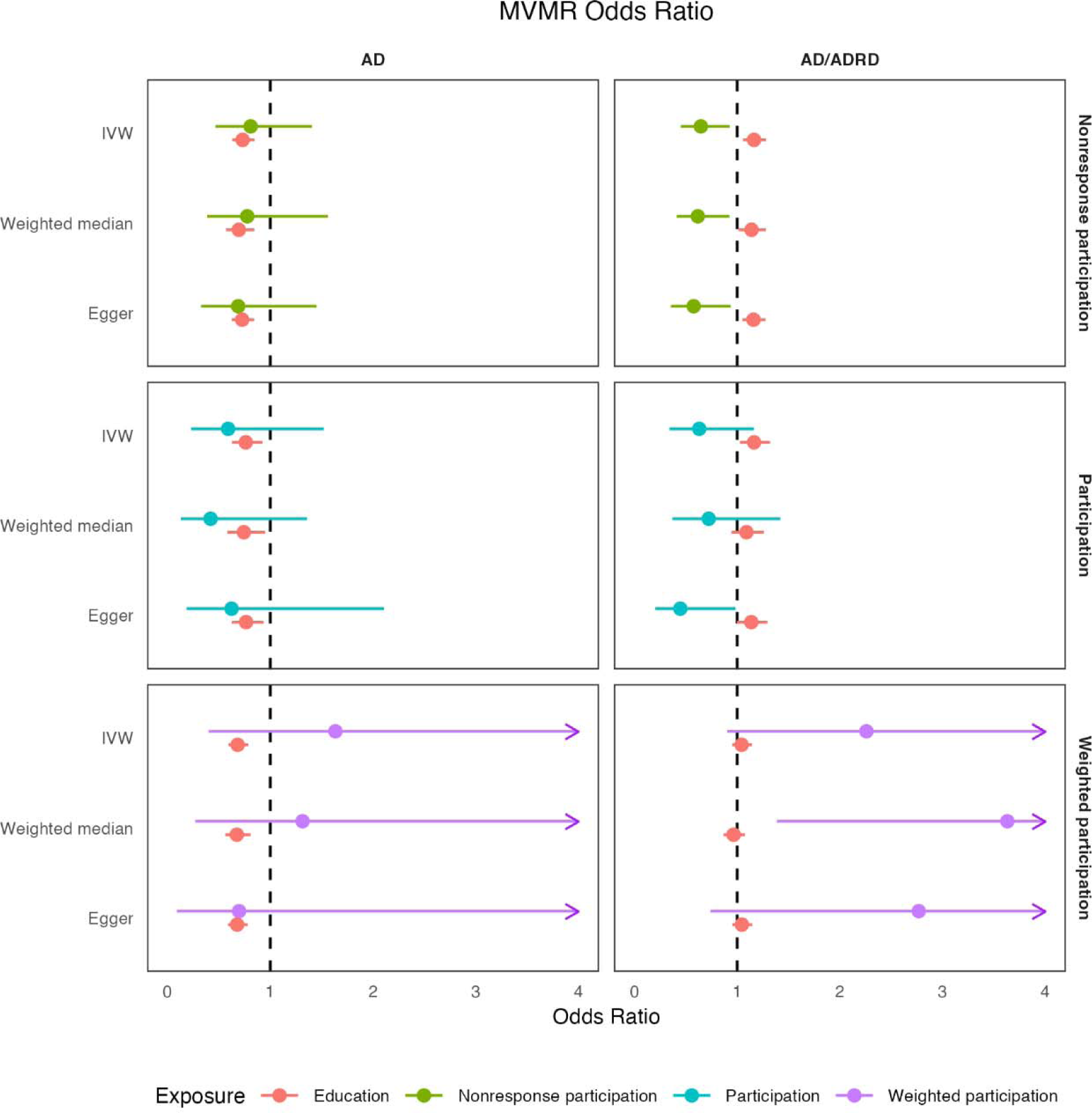
MVMR odds ratio of the association between participation measures and education for AD and AD/ADRD. Forest plot examining MVMR associations between education, three distinct participation measures and clinical AD versus AD/ADRD. Each row represents a different participation measure and arrows represent CI intervals that continue past the x axis.

When adjusting for participation in the MHQ in MVMR, an additional year of education increased the odds of AD/ADRD (OR_IVW_ 95% CI= 1.16 [1.03, 1.32], p = 0.02; Figure 4 and Supplementary Table S3). However, this was not robust, and our analysis suffered from weak instruments (conditional F statistic for education= 3.73 and participation= 2.48). This finding was supported when adjusting for nonresponse participation; years of education increased odds of AD/ADRD (OR_IVW_ 95% CI= 1.16 [1.06, 1.28], p = 0.002; Figure 4 and Supplementary Table S3) with robust findings, though instrument strength remained weak (conditional F statistic for education= 7.42 and nonresponse participation= 2.79). When adjusting for weighted participation, our results were non-significant (Supplementary Table S3).

The SNPS used as IVs and their harmonized effects are shown in Supplementary Tables S4 to S19. The results from Radial MR analysis are displayed in Supplementary Figures 6-9.

## Discussion

This study used genetic correlations, univariable MR, and multivariable MR to evaluate the causal relationships between education, participation, and AD risk. Higher education was associated with a reduced risk of AD but an increased risk of AD/ADRD with evidence of heterogeneity, indicating that these effects could have alternative explanation (e.g. horizontal pleiotropy). Participation was also associated with a reduced risk of AD and AD/ADRD. When combining education and participation into a multivariable framework, we found that the relationship between education and AD was unlikely to be due to participation.

Education exhibited the strongest positive correlation with weighted participation, followed by participation and nonresponse participation. There were also significant negative correlations between educational attainment and AD, supporting consistent findings on education’s protective effect on AD (13,14). Higher levels of education seem to play a crucial role in maintaining cognitive function across an extended lifespan (36). Educational attainment significantly contributes to cognitive reserve, an individual’s ability to perform tasks and solve problems, even in the presence of amyloid pathology (37). Specifically, studies have shown that individuals with higher educational attainment exhibited better cognitive abilities among carriers of PSEN1 and E280A (mutations that predispose individuals to early-onset AD) (38). Education is thought to contribute to cognitive reserve by increasing synaptic density in the neocortical association cortex (39).

Across both AD and AD/ARRD datasets, participation in the MHQ was associated with a reduced risk of AD. This finding was supported by a significant negative genetic correlation between participation and AD. Participation bias in studies could lead to a non-representative population, which can produce results that are not generalizable to other populations (17–19). Factors influencing participation in clinical research include lower socioeconomic status (lower wages, lower quality neighborhoods, higher unemployment, and household overcrowding) and lower educational achievement, which are risk factors for AD and are closely linked to the risk of cognitive impairment (40,41). Furthermore, weighted participation and participation exhibited strong genetic correlations, suggesting that similar genetic factors drive participation, whether in a mental health questionnaire or other aspects of health, lifestyle, and education (18).

The bidirectional analysis in both the AD and AD/ADRD datasets provided evidence supporting an apparent causal effect of genetic liability to AD on participation. This suggests that AD pathogenesis may influence participation in cohort studies in mid-life, which aligns with the development of AD pathology at least 15 years before diagnosis (1). Genetic liability to AD/ADRD was associated with a small decrease in participation but a small increase in weighted participation. This discrepancy in the weighted participation measure analysis can be attributed to the use of proxy measures for both AD and participation, which may lead to inaccuracies. Nevertheless, the effect of AD on participation can largely be explained through education; lower educational levels are linked to a decreased likelihood of participation among individuals with AD (42).

To distinguish the individual impacts of participation and education on AD, we conducted MVMR. Our MVMR analysis, adjusting for participation, revealed that the effect of education on AD remained substantial, suggesting that the effect of years of education on AD found using Mendelian randomization studies is unlikely to be due to participation bias but is likely mediated by other mechanisms. Given the mounting evidence supporting the effect of education on risk of AD, it becomes crucial to implement robust public policies encouraging sustained education, and to discover the molecular, familial, and societal mechanisms that mediate these effects. Research indicates that a population-wide preventive approach for AD could involve focusing on the duration of mandatory schooling and promoting cognitive engagement in the elderly as active aging (based on social and intellectual advanced activities of daily living) may be associated with cognitive performance (14,43,44).

Discrepancies in results emerged between the AD and AD/ADRD datasets. Notably, the univariate MR analysis indicated that education had a protective effect on AD but a risk-increasing effect for AD/ADRD. This effect is likely due to survival bias and reporting bias (45). The AD/ADRD cases in the Bellenguez dataset are largely comprised of individuals who reported parental dementia, and individuals reporting parental AD diagnoses may have parents with less genetic risk for other diseases, such as cardiovascular disease and cancer, allowing them to live longer. More educated parents will be older when they have kids and will have more time to develop AD, which is supported by positive genetic correlations between age at first birth and educational attainment (46). Additionally, only individuals aware of their parent’s health typically report parental AD diagnoses (45). While efforts can be made to mitigate AD biases in GWAX, such as controlling for parental age and vital status, the ultimate solution lies in enhancing the quality of GWAX analysis. These discrepancies highlight the importance of using GWAS datasets containing clinical or neuropathological defined cases over family history-based proxy phenotypes for MR studies (47).

Strengths of our study include using complementary measures of participation, which allowed us to see the effectiveness of different methods; a bidirectional MR approach to confirm the direction of causality; comparison of clinical versus proxy datasets; and multiple sensitivity analyses to confirm the robustness of the results. Nevertheless, our study findings come with several limitations. Firstly, while F-statistics for the univariable analyses exceed the standard threshold of 10, the conditional F statistics in our MVMR analyses were below 10. This indicates that causal estimates in the MVMR analyses may be affected weak instrument bias, which can be attributed to the educational dataset possessing more statistical power than the participation datasets. There has been a lack of highly powered participation datasets and future work can focus on implementing methods to address weak instrument bias (48). Secondly, the datasets exclusively include individuals of European ancestry, limiting the generalizability of our results to the broader population. Thirdly, biases might arise when there is a proportion of sample overlap between datasets (49).

In conclusion, we found that the Mendelian randomization estimates of the effect of years of education on AD are unlikely to be due to participation bias. Education increased the odds of AD/ADRD in our GWAX dataset, likely attributed to survival bias and reporting bias, highlighting the importance of utilizing clinical case-control AD GWAS in MR analyses. These findings contribute to the accumulating evidence supporting the protective role of education against AD. We urgently need to discover the mechanisms that explain these effects and develop effective interventions to reduce the incidence of AD in the population.

## Supporting information

Strobe MR checklist

## Acknowledgements

The authors thank the investigators of the GWAS’s used in this study and the research participants.

## Data Availability

Harmonized data is available in supplementary tables; https://github.com/AndrewsLabUCSF/Aadrita-AD-participation-education

Original summary statistics can be found here:

Kunkle AD- https://www.niagads.org/datasets/ng00075

Bellenguez AD/ADRD- https://www.ebi.ac.uk/gwas/publications/35379992

Education- http://www.thessgac.org/data

Participation in MHQ- https://www.ebi.ac.uk/gwas/publications/33563987

Weighted participation- https://www.ebi.ac.uk/gwas/publications/37106081

Nonresponse participation- https://www.ebi.ac.uk/gwas/publications/37386106

---

**Supplementary Table 1: Description of GWAS datasets used in this study.**

**Supplementary Table 2: Summary of MVMR results with AD**

**Supplementary Table 3: Summary of MVMR results with AD/ADRD**

**Supplementary Table 4: Harmonized data for AD onto participation bidirectional analysis**

**Supplementary Table 5: Harmonized data for AD onto nonresponse participation bidirectional analysis**

**Supplementary Table 6: Harmonized data for AD onto education bidirectional analysis**

**Supplementary Table 7: Harmonized data for AD onto weighted participation bidirectional analysis**

**Supplementary Table 8: Harmonized data for AD/ADRD onto participation bidirectional analysis**

**Supplementary Table 9: Harmonized data for AD/ADRD onto nonresponse participation bidirectional analysis**

**Supplementary Table 10: Harmonized data for AD/ADRD onto weighted participation bidirectional analysis**

**Supplementary Table 11: Harmonized data for AD/ADRD onto education bidirectional analysis**

**Supplementary Table 12: Harmonized data for participation onto AD univariate MR analysis**

**Supplementary Table 13: Harmonized data for nonresponse participation onto AD univariate MR analysis**

**Supplementary Table 14: Harmonized data for weighted participation onto AD univariate MR analysis**

**Supplementary Table 15: Harmonized data for education onto AD univariate MR analysis**

**Supplementary Table 16: Harmonized data for participation onto AD/ADRD univariate MR**

**Supplementary Table 17: Harmonized data for nonresponse participation onto AD/ADRD**

**Supplementary Table 18: Harmonized data for weighted participation onto AD/ADRD univariate MR analysis**

**Supplementary Table 19: Harmonized data for education onto AD/ADRD univariate MR analysis**

**Supplementary Fig 1. Directed acyclic graph (DAG) illustrating collider bias between our traits of interest.**

**Supplementary Fig 2. Univariate MR Scatter Plots for AD**

**Supplementary Fig 3. Univariate MR Scatter Plots for AD/ADRD**

**Supplemental Fig 4. Bidirectional analyses for AD onto education and participation measures**

**Supplemental Fig 5. Bidirectional analyses for AD/ADRD onto education and participation measures**

**Supplementary Fig 6. Radial MVMR for AD**

**Supplementary Fig 7. Radial MVMR for AD/ADRD**

**Supplementary Fig 8. Radial MR plots for AD**

**Supplementary Fig 9. Radial MR plots for AD/ADRD**

**Supplementary Methods**

**STROBE-MR Checklist**

