## Supplementary Information for "Disentangling the causal effects of education and participation bias on Alzheimer’s disease using Mendelian Randomization"

**Table of Contents**

Supplementary Table S1. Description of GWAS datasets used in this study	3

Supplementary Table S2. Summary of MVMR results with AD	5

Supplementary Table S3. Summary of MVMR results with AD/ADRD	6

Supplementary Fig 1. Directed acyclic graph (DAG) illustrating collider bias between our traits of interest	7

Supplementary Fig 2. Univariate MR Scatter Plots for AD	8

Supplementary Fig 3. Univariate MR Scatter Plots for AD/ADRD	9

Supplemental Fig 4. Bidirectional analyses for AD onto education and participation measures	10

Supplemental Fig 5. Bidirectional analyses for AD/ADRD onto education and participation measures	11

Supplementary Fig 6. Radial MVMR for AD	12

Supplementary Fig 7. Radial MVMR for AD/ADRD	13

Supplementary Fig 8. Radial MR plots for AD	14

Supplementary Fig 9. Radial MR plots for AD/ADRD	15

Supplementary Methods	16

References	16

**Supplementary Table S1. Description of GWAS datasets used in this study.**

| Study | Trait | Details | Cohort/consortium | N | Mean age |
| --- | --- | --- | --- | --- | --- |
| Kunkle et al, 2019 | Late-onset Alzheimer’s disease | The study was broken up into 3 stages of analysis: the first was a meta-analysis of non-Hispanic Whites (NHW) (46 total datasets; n = 21,982 cases, 41,944 cognitively normal controls). The second stage involved replication with existing custom genotyping chip and the third stage had stage 3A (n = 11,666) and stage 3B (n = 30,511). Alzheimer’s disease patients had a clinical diagnosis of AD or were autopsy documented cases. The total sample size had 35,274 clinical and autopsy cases and 59,163 controls. | IGAP | 94,437 | 58-86 |
| Bellenguez et al, 2022 | Proxy AD | The study consisted of two stages: the first stage had the European Alzheimer & Dementia Biobank (EADB), which consists of various European GWAS consortia, and was meta-analyzed with a proxy-AD UKB dataset. There were 39,106 clinically diagnosed AD cases, 46,828 proxy AD cases from the UKBB, and 401,577 controls. The sample was filtered by standard quality control measures. The second stage had ADGC, FinnGen, and CHARGE consortia and contained 25,392 AD cases and 276,086 controls. | EADB consortium Band the UK Biobank | 788, 989 | N/A |
| Okbay et al, 2022 | Education | A meta-analysis was performed on three different sets of summary statistics including Lee et al. (2018) and 23andMe (n= 2,272,216) and UK Biobank association results (n=441,121). All participants were of European ancestry who passed quality control measures set by the cohort (apart from 23andMe and UKB results that used the EasyQC R package). Data was collected independently from multiple cohorts and data with less than seven years of schooling were eliminated as outliers from the analysis. | UK Biobank | 3,037,499 | 30 years and older |
| Tyrrell et al, 2021 | Participation in MHQ | Participation was defined as a binary variable with 0 representing invited but not accepted and 1 representing invited and accepted. The UKB first sent out an invitation email and sent a reminder email to the nonresponders and partial responders and after 4 months, sent a last invitation email. The researchers used individuals who were defined as European ancestry using principal component analyses. The quality control process involved excluding variants that had an imputation quality (INFO) <0.3 or minor allele frequency (MAF) <0.1%. Lead SNPs consisted of the smallest P value and locus boundaries were set within a 0.5 Mb distance from the lead SNP. The relationship between participation demographics and participation measures were calculated using logistic regression. | UK Biobank | 294,787 | 55-56 |
| Schoeler et al, 2021 | Weighted participation | Only UK Biobank participants who passed standard GWA analysis quality control measures were included. This model adjusted for nonresponse by giving greater weight to overrepresented and underrepresented individuals, thus creating a more representative pseudo-population that mimics the Health Survey England (reference sample). All analyses were adjusted for batch, principal components (PC1-5), age and sex. The sample was filtered by geographic regions in order to match the geographical regions in the reference sample. All participants provided written informed consent to participate. | UK Biobank, HSE, UK Census | 94,643 – 102,215 | 40-69 |
| Mignogna et al, 2023 | Nonresponse participation | Participants passed standard quality control measures and individuals who withdrew from the study without filling out the touchscreen survey were excluded from the analysis (N=231). The “I don’t know” phenotype was also analyzed in Add Health which is a nationally representative sample of US adolescents enrolled in grades 7 through 12 during the 1994-1995 school year. Exploratory factor analysis was used to generate estimated factor scores. | UK Biobank, Add Health | 360,628 | 40-69 in UKB and 29 in Add Health |

**Supplementary Table S2. Summary of MVMR results with AD**

|  | | SNP | F stats |  | Fixed- effects IVW | |  | MR-Egger | |  | Weighted median | |  |  | | Cochran’s Q test | |
| --- | --- | --- | --- | --- | --- | --- | --- | --- | --- | --- | --- | --- | --- | --- | --- | --- | --- |
| Exposure | Outcome |  |  |  | Beta (se) | P value |  | Beta (se) | P value |  | Beta (se) | P value |  | |  | | Q p value |
| Education | AD | 461 | 3.41 |  | -0.27 (0.09) | 0.006 |  | -0.26 (0.1) | 0.008 |  | -0.29 (0.12) | 0.01 |  | |  | | 0.0001 |
| Participation | AD | 11 | 2.41 |  | -0.52 (0.48) | 0.27 |  | -0.47 (0.62) | 0.44 |  | -0.87 (0.59) | 0.14 |  | |  | |  |
| Education | AD | 461 | 10.8 |  | -0.38 (0.07) | 1.22e-07 |  | -0.39  (0.07) | 7.7e-08 |  | -0.39 (0.09) | 2.22e-05 |  | |  | | 0.0003 |
| Weighted participation | AD | 4 | 1.4 |  | 0.49 (0.71) | 4.9e-01 |  | -0.36 (1.04) | 7.27e-01 |  | 0.27 (0.8) | 7.35e-01 |  | |  | |  |
| Education | AD | 293 | 7.52 |  | -0.31 (0.07) | 3.54e-05 |  | -0.32 (0.07) | 2.92e-05 |  | -0.36 (0.1) | 2.89e-04 |  | |  | | 4.08e-04 |
| Nonresponse participation | AD | 14 | 2.82 |  | -0.21 (0.28) | 0.45 |  | -0.37 (0.38) | 0.32 |  | -0.25 (0.35) | 0.47 |  | |  | |  |

**Supplementary Table S3. Summary of MVMR results with AD/ADRD**

|  | | SNP | F stats |  | Fixed- effects IVW | |  | MR-Egger | |  | Weighted median | |  |  | | Cochran’s Q test | |
| --- | --- | --- | --- | --- | --- | --- | --- | --- | --- | --- | --- | --- | --- | --- | --- | --- | --- |
| Exposure | Outcome |  |  |  | Beta (se) | P value |  | Beta (se) | P value |  | Beta (se) | P value |  | |  | | Q p value |
| Education | AD/ADRD | 11 | 3.73 |  | 0.15 (0.06) | 0.017 |  | 0.12 (0.06) | 0.05 |  | 0.08 (0.07) | 0.23 |  | |  | | 2.15e-17 |
| Participation | AD/ADRD | 464 | 2.4 |  | -0.46 (0.31) | 0.14 |  | -0.8 (0.4) | 0.04 |  | -0.32 (0.34) | 0.34 |  | |  | |  |
| Education | AD/ADRD | 463 | 10.6 |  | 0.04 (0.04) | 0.36 |  | 0.04 (0.04) | 0.34 |  | -0.03 (0.05) | 0.13 |  | |  | | 5.6e-17 |
| Weighted participation | AD/ADRD | 4 | 1.43 |  | 0.81 (0.46) | 0.08 |  | 1.02 (0.67) | 0.13 |  | 1.29 (0.49) | 0.008 |  | |  | |  |
| Education | AD/ADRD | 468 | 7.42 |  | 0.15 (0.04) | 0.002 |  | 0.147 (0.04) | 0.003 |  | 0.13 (0.05) | 0.02 |  | |  | | 1.43e-16 |
| Nonresponse participation | AD/ADRD | 10 | 2.79 |  | -0.43 (0.18) | 0.01 |  | -0.55 (0.24) | 0.02 |  | -0.48 (0.2) | 0.01 |  | |  | |  |

**Supplementary Fig 1. Directed acyclic graph (DAG) illustrating collider bias between our traits of interest.**

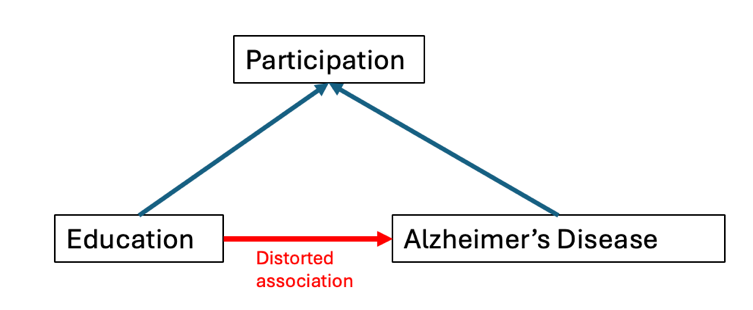

**Supplementary Fig 2. Univariate MR Scatter Plots for AD**
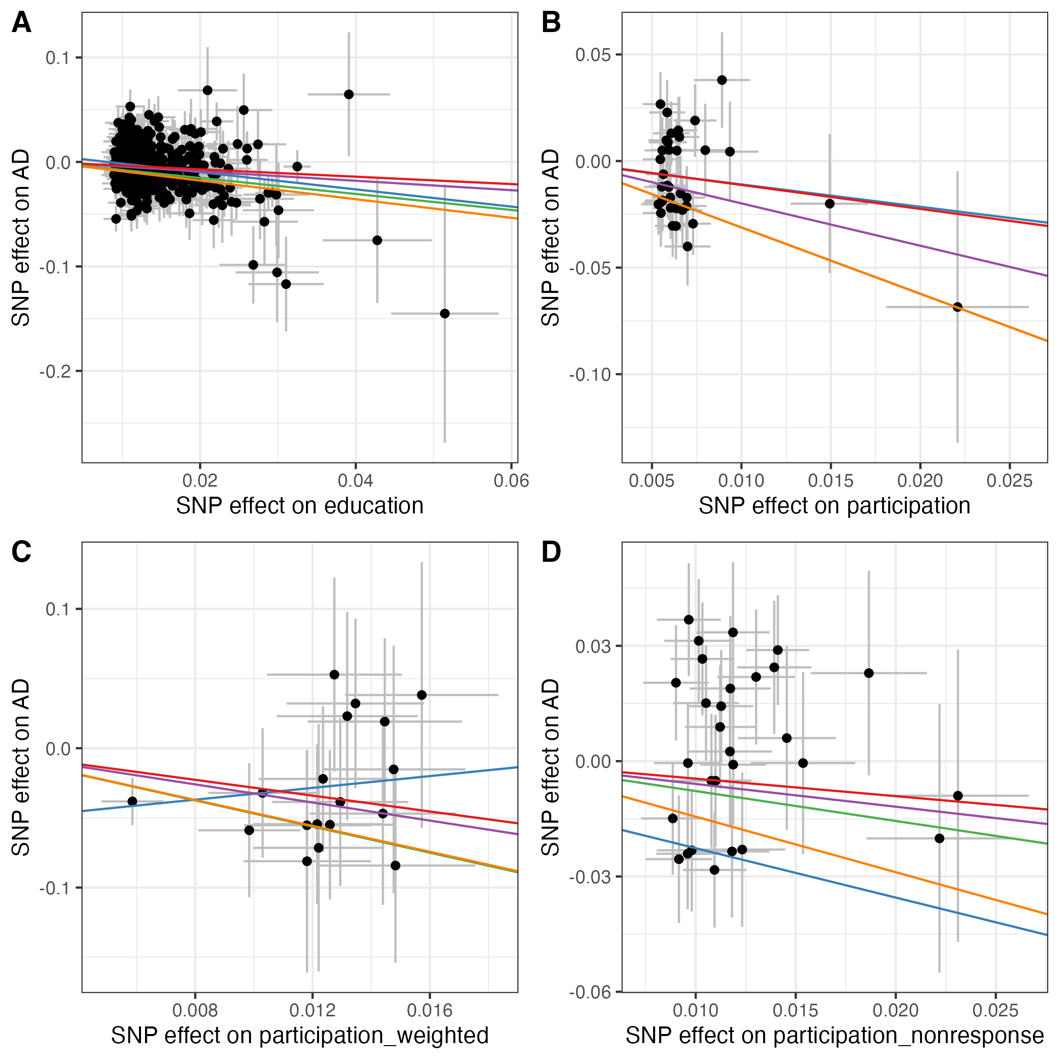

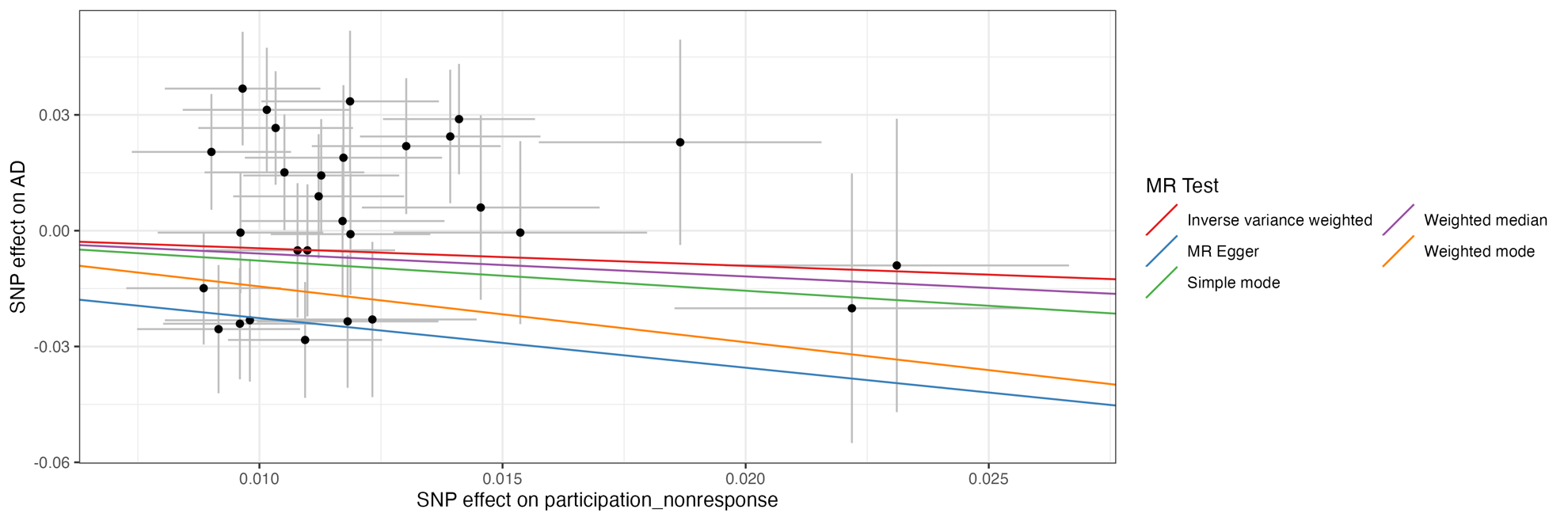

Scatter plot depicting the relationship between SNP effects on (A) education (B) participation (C) weighted participation (D) nonresponse participation and SNP effects on AD. The slope of the lines represents the estimated causal effect for each method.

**Supplementary Fig 3. Univariate MR Scatter Plots for AD/ADRD**

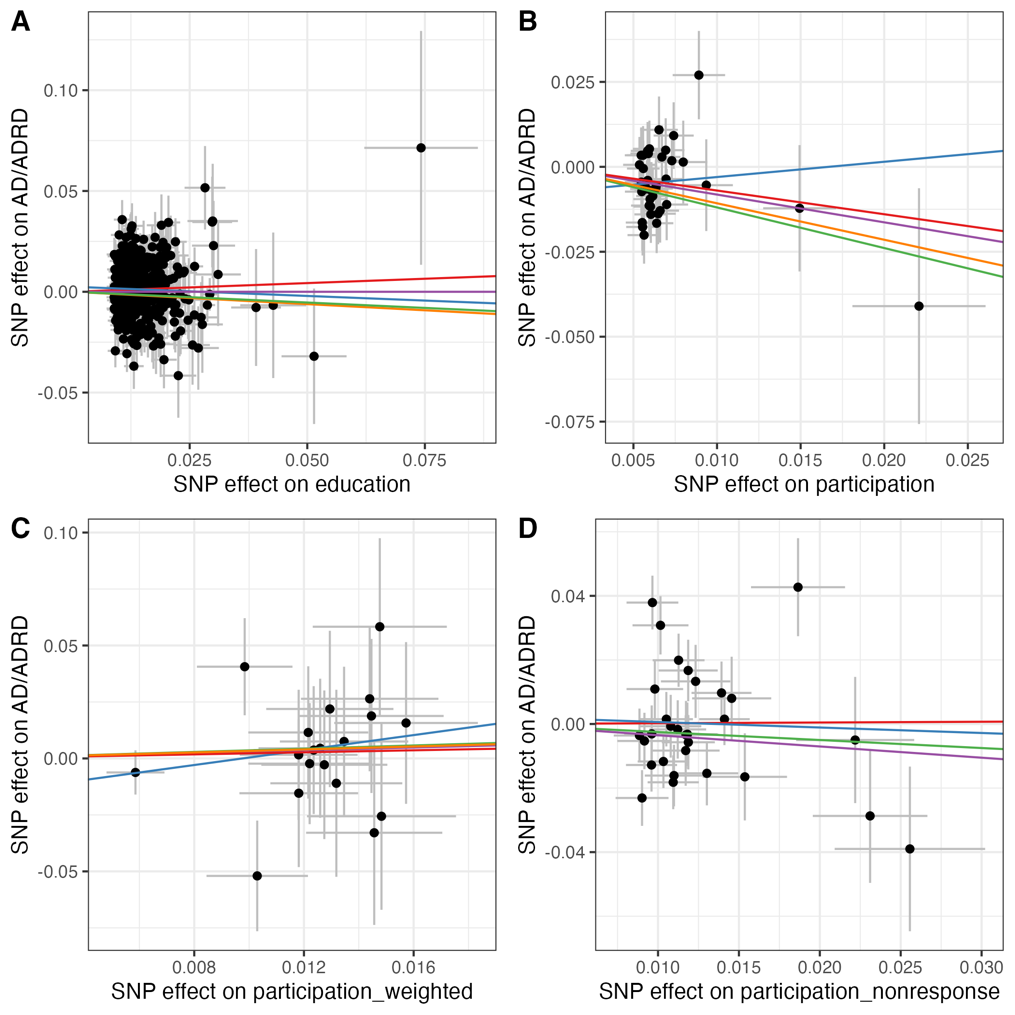

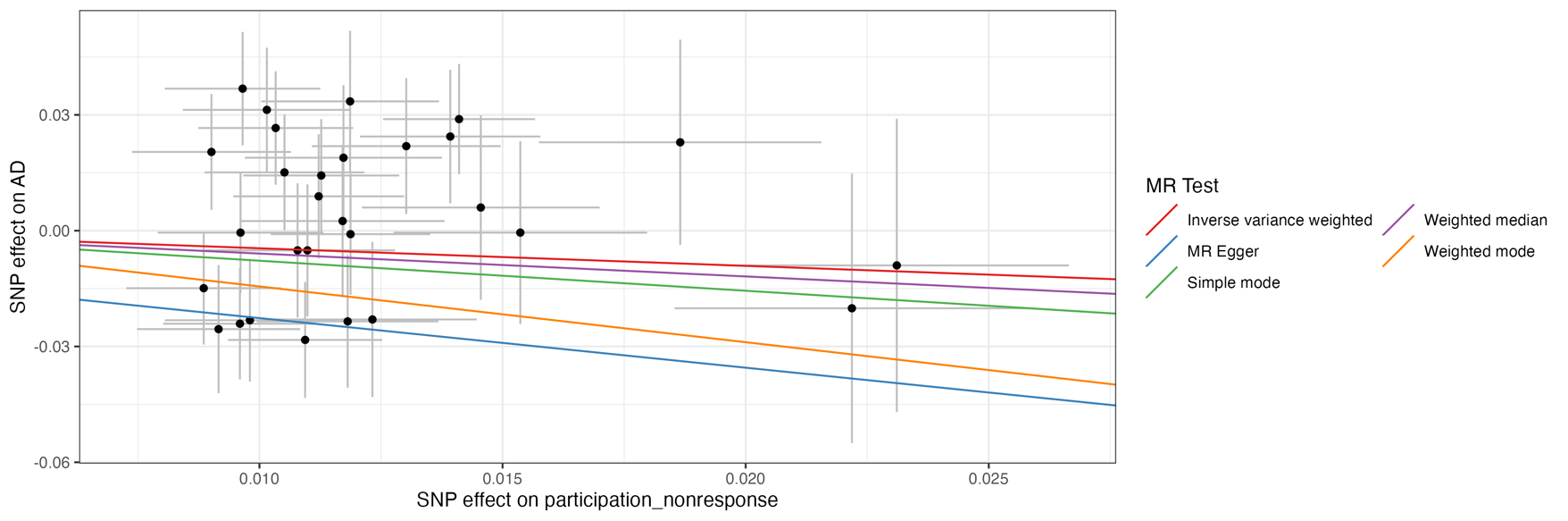

Scatter plot depicting the relationship between SNP effects on (A) education (B) participation (C) weighted participation (D) nonresponse participation and SNP effects on AD/ADRD. The slope of the lines represents the estimated causal effect for each method.

**Supplemental Fig 4. Bidirectional analyses for AD onto education and participation measures**

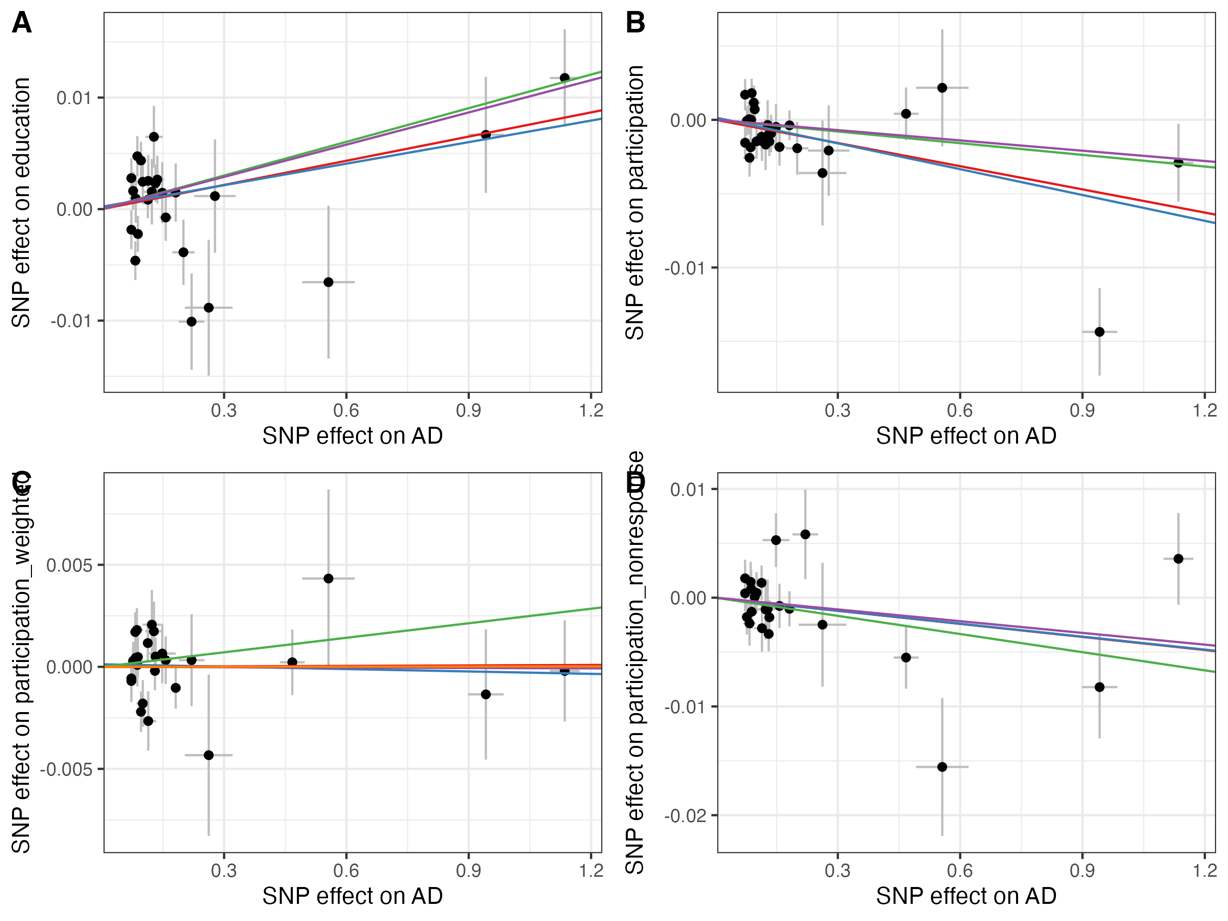

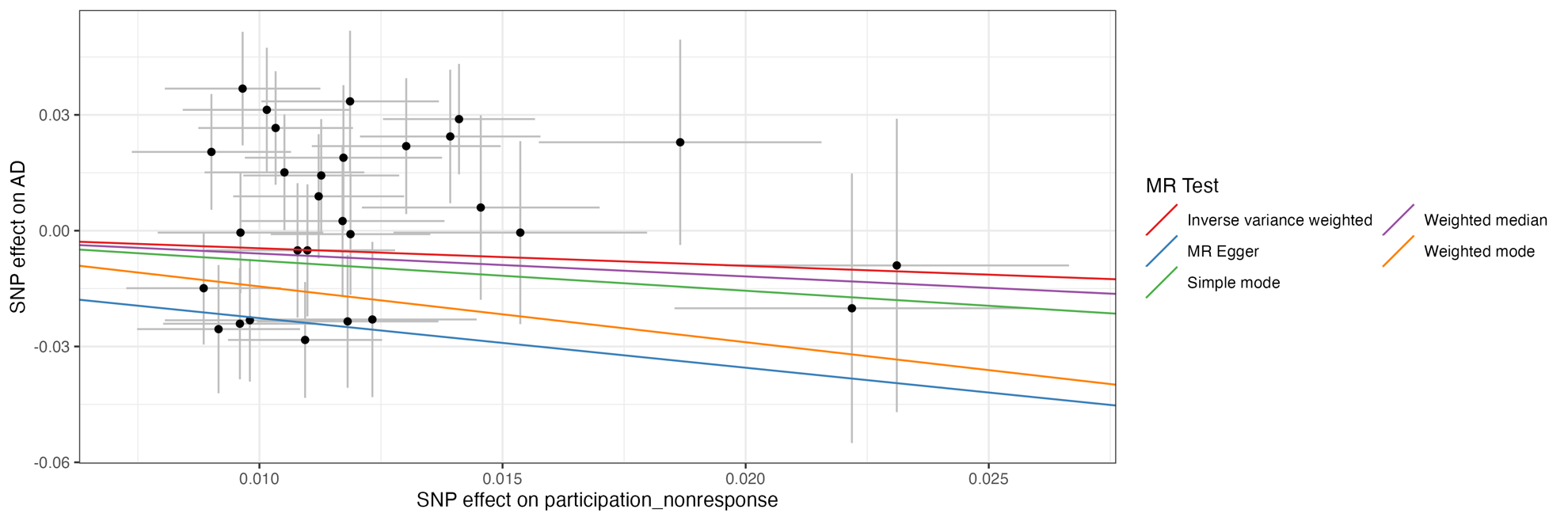

Scatter plot depicting the relationship between SNP effects on AD and SNP effects on (A) education (B) participation (C) weighted participation and (D) nonresponse participation. The slope of the lines represents the estimated causal effect for each method.

**Supplemental Fig 5. Bidirectional analyses for AD/ADRD onto education and participation measures**

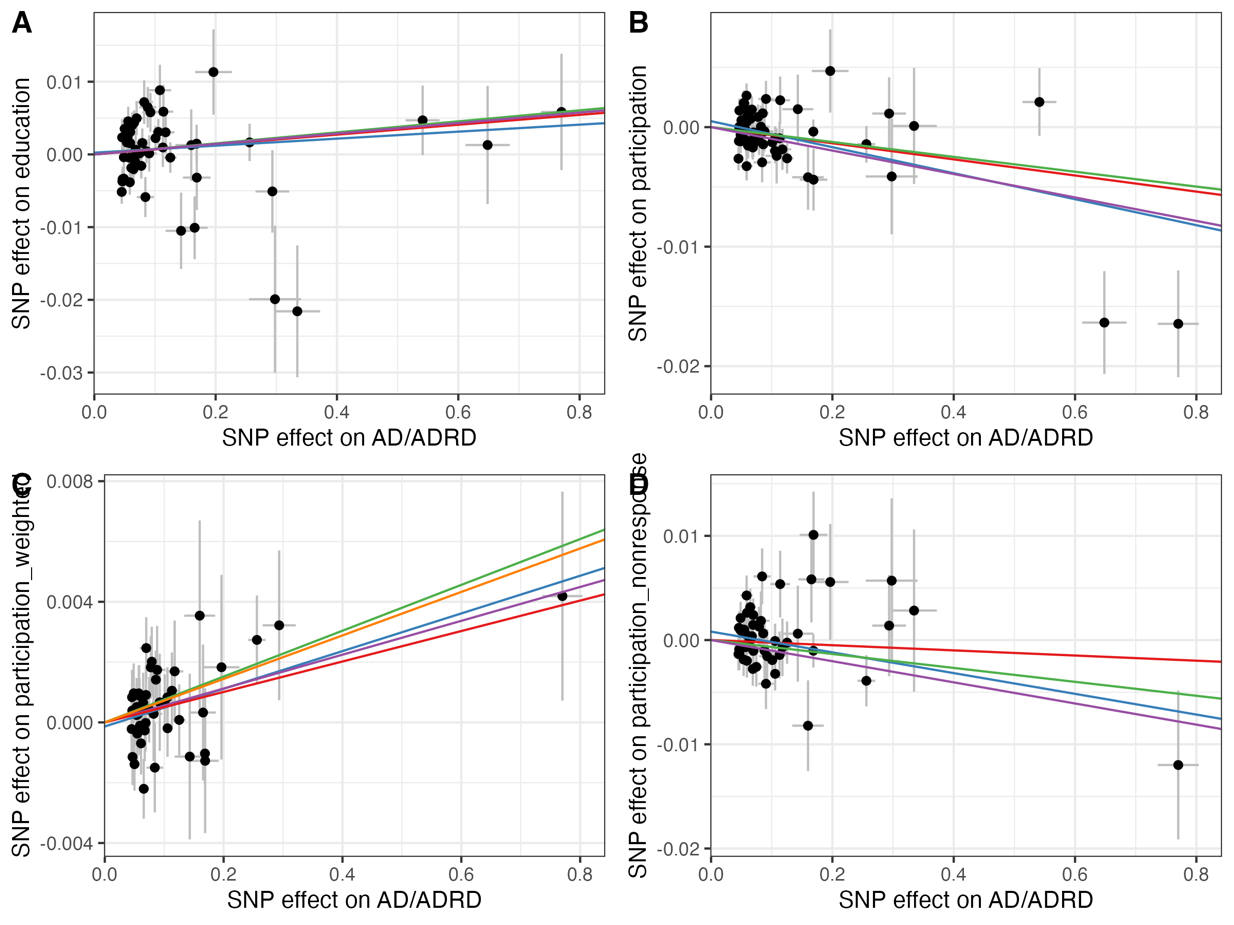

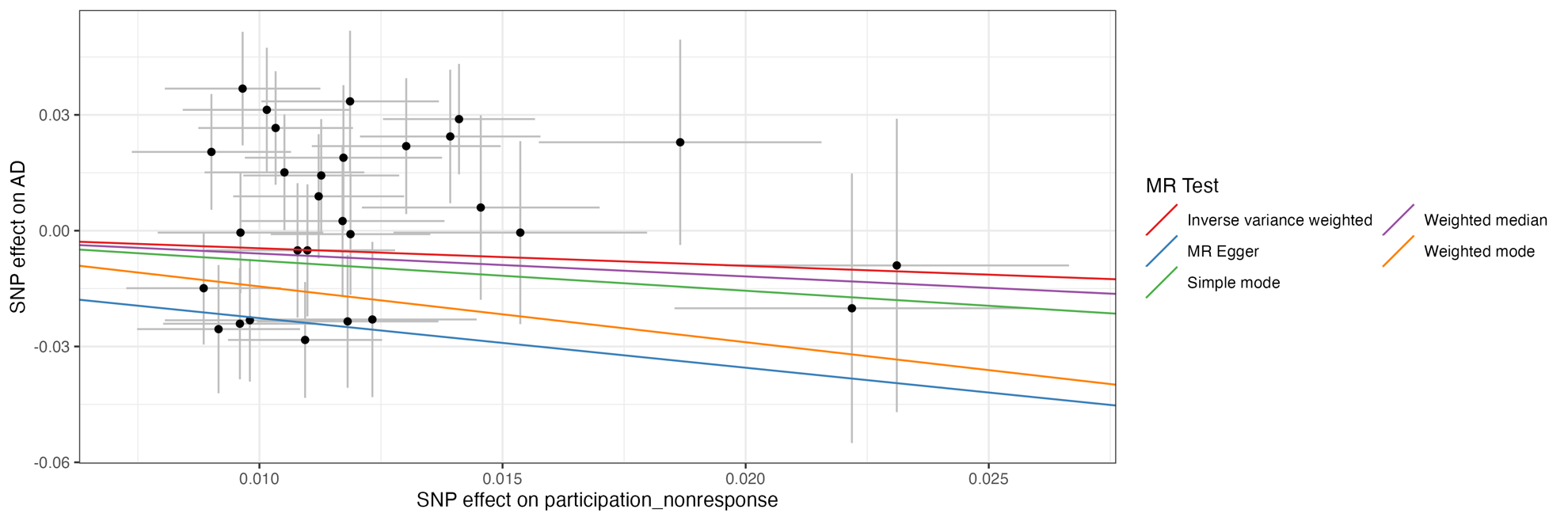

Scatter plot depicting the relationship between SNP effects on AD/ADRD and SNP effects on (A) education (B) participation (C) weighted participation and (D) nonresponse participation. The slope of the lines represents the estimated causal effect for each method.

**Supplementary Fig 6. Radial MVMR for AD**

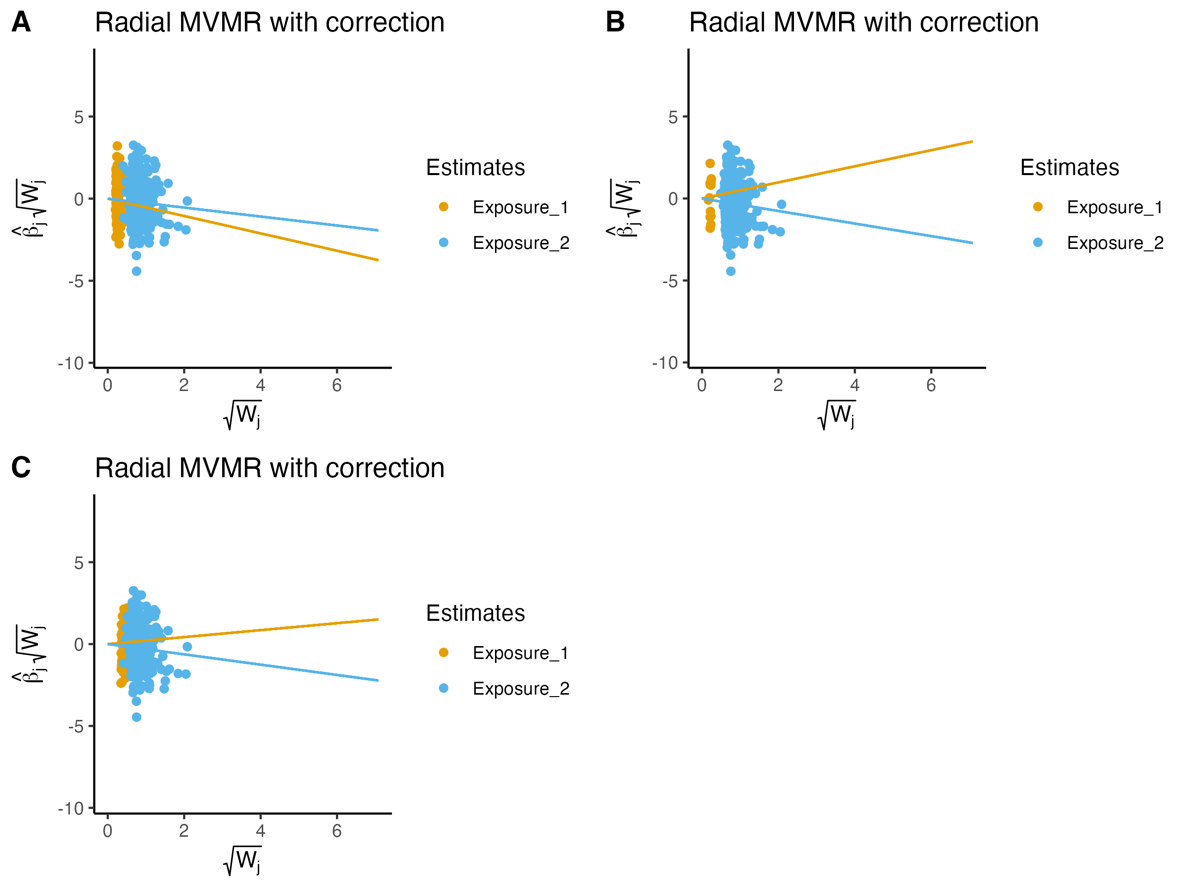

Radial MVMR plots showing the causal effect of (A) Participation- Education- AD MVMR (B) Weighted participation- Education- AD MVMR (C) Nonresponse- Education- AD MVMR on AD as a function of the ratio estimate and weighting corresponding to each SNP. Outliers were removed.

**Supplementary Fig 7. Radial MVMR for AD/ADRD**

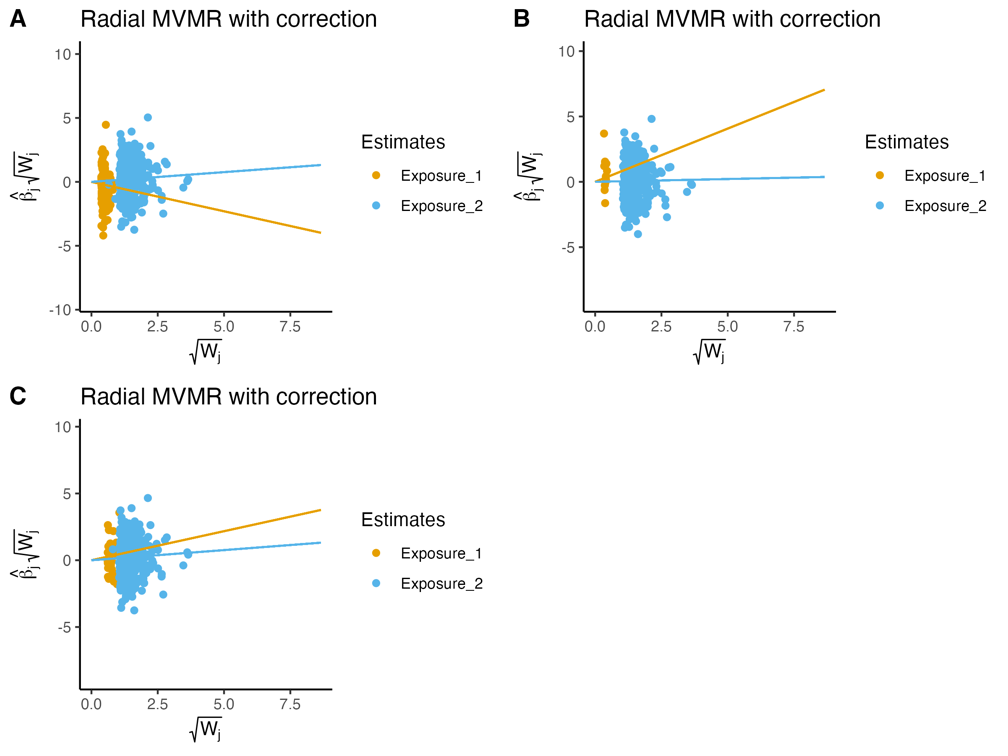

Radial MVMR plots showing the causal effect of (A) Participation- Education- AD MVMR (B) Weighted participation- Education- AD MVMR (C) Nonresponse- Education- AD MVMR on AD/ADRD as a function of the ratio estimate and weighting corresponding to each SNP. Outliers were removed.

**Supplementary Fig 8. Radial MR plots for AD**

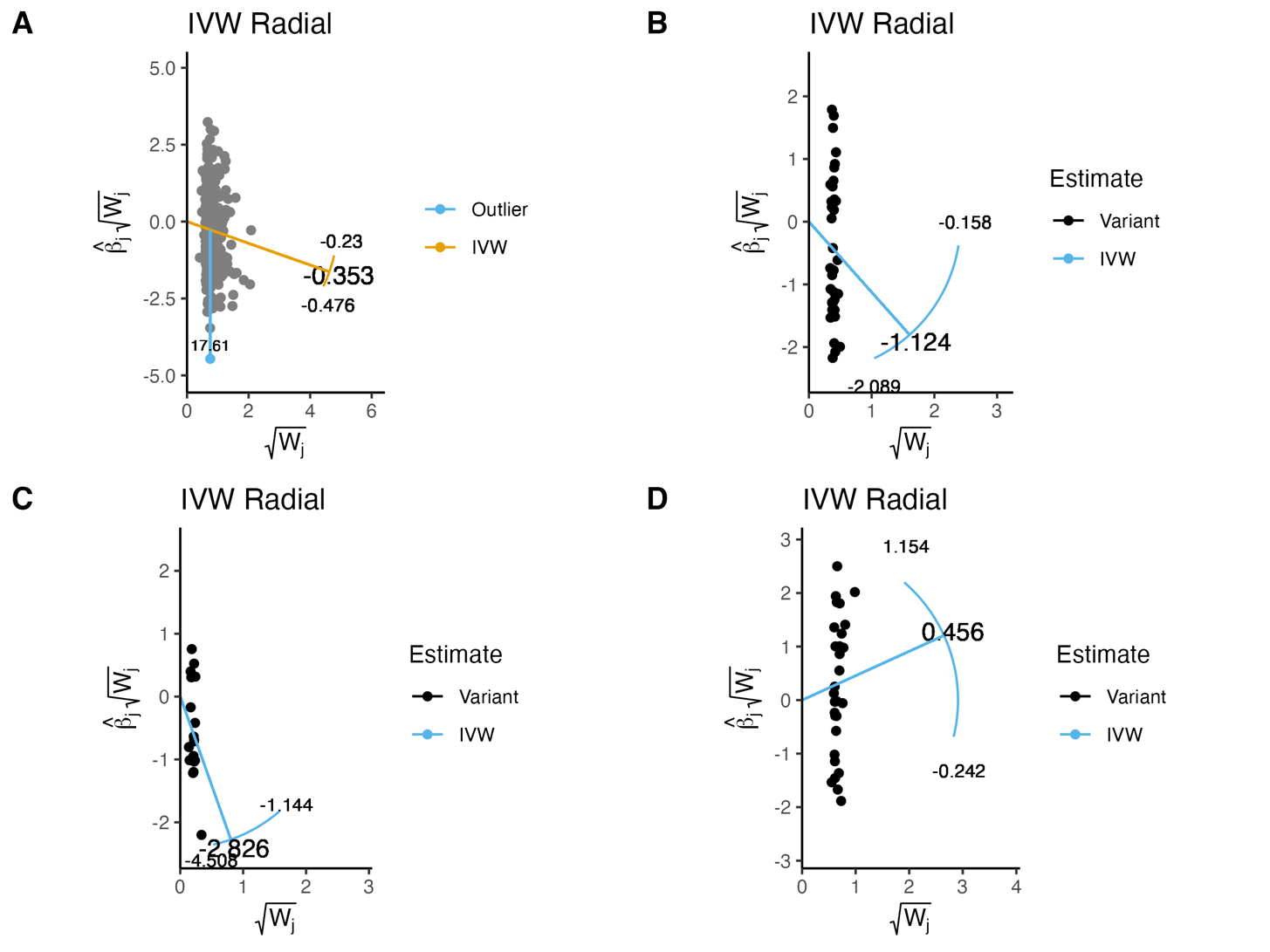

Radial plots showing the regression line (blue line) and radial causal estimate for the inverse variance weighted method for (A) education (B) participation (C) weighted participation (D) nonresponse participation.

**Supplementary Fig 9. Radial MR plots for AD/ADRD**

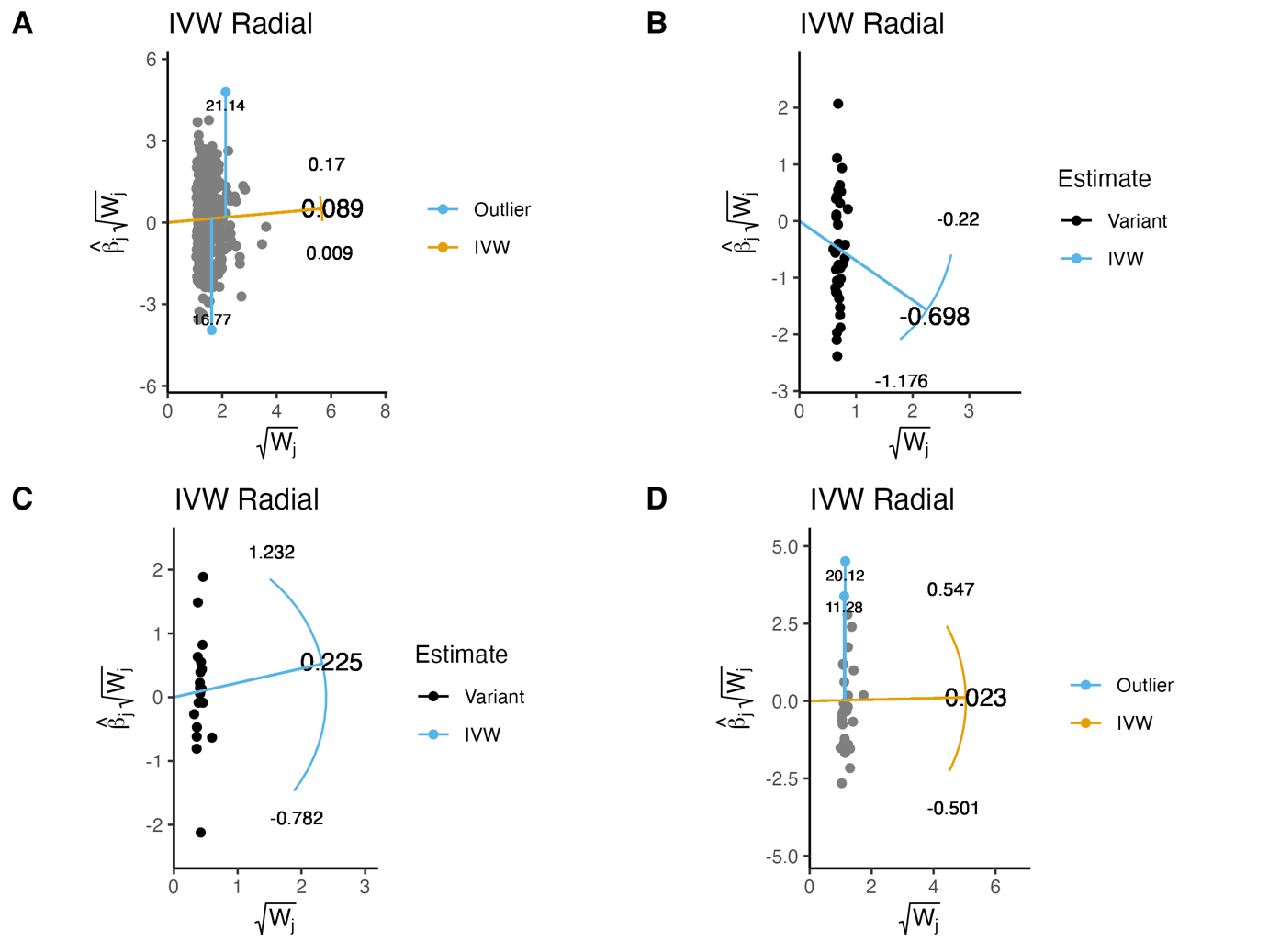

Radial plots showing the regression line (blue line) and radial causal estimate for the inverse variance weighted method for (A) education (B) participation (C) weighted participation (D) nonresponse participation.

**Supplementary Methods**

Diagnostic tests: Diagnostic tests for MR are important to assess the validity of the instrumental variable assumptions and the reliability of causal inference. These include Cochran’s Q test for heterogeneity and the MR Egger regression intercept for horizontal pleiotropy. Heterogeneity refers to differences in causal estimates between the exposure and outcome and significant heterogeneity may indicate that the instrumental variable assumptions are violated. Horizontal pleiotropy occurs when genetic variants have direct effects on the outcome independent of the exposure, with a non-zero intercept in the Egger regression suggesting the presence of directional pleiotropy (1).

Genetic correlations: Positive genetic correlations indicate that two traits share genetic factors that act in the same direction. In other words, individuals with genetic variants associated with higher values for one trait are more likely to have genetic variants associated with higher values for the other trait, and vice versa. Negative genetic correlations, on the other hand, suggest that two traits share genetic factors that act in opposite directions (2).
